# Target trial emulation of physical activity and cardiovascular disease risk: What is impact of the exposure assessment method?

**DOI:** 10.1101/2025.11.02.25339322

**Authors:** Matthew Ahmadi, Borja del Pozo Cruz, Raaj Biswas, Nicholas Koemel, Armando Teixeira-Pinto, Dot Dumuid, Marla Beauchamp, Joanne McVeigh, Mark Hamer, Emmanuel Stamatakis

**Affiliations:** Mackenzie Wearables Research Hub; Charles Perkins Centre, University of Sydney; School of Health Sciences, Faculty of Medicine and Health, University of Sydney; Department of Sport Sciences, Faculty of Medicine, Health, and Sports, Universidad Europea de Madrid, Villaviciosa de Odón, Madrid, 28670, Spain; Snow Vision Accelerator, University of Sydney; Alliance for Research in Exercise, Nutrition and Activity, Allied Health & Human Performance, University of South Australia, Adelaide, South Australia, Australia; School of Rehabilitation Science, McMaster University, Hamilton, ON, Canada; Curtin School of Allied Health, Curtin University, Perth, Western Australia; Institute of Sport Exercise and Health, Division of Surgery and Interventional Sciences, University College London, United Kingdom

**Author notes:** **Corresponding author:** Emmanuel Stamatakis, **Address for correspondence:** Hub D17, Charles Perkins Centre L6 West, the University of Sydney, New South Wales, Australia. Joint first author (Matthew Ahmadi, Borja del Pozo Cruz).

## Abstract

**Background:** Target trial emulation (TTE) designs provide a framework for strengthening causal inference in observational research, but it is unknown how vulnerable they are to substantial error when the emulated intervention (exposure) measurement is imprecise. In physical activity epidemiology specifically, correcting for confounding via TTE designs but not addressing the large measurement errors arising from self-reports (e.g. questionnaires, which typically capture partial behavioural accounts with very low precision) creates uncertainty about possible dominance of type 2 error biases arising from such novel designs. High-resolution wearables-based methods capture most movement, providing an assessment of physical activity behaviour with substantially less, empirically verified, measurement error. No study has examined how physical activity measurement method influences causal inference in TTE studies.

**Objectives:** We applied TTE methodology to sub-samples of the UK Biobank cohort with repeat exposure measurements, to compare the effects of an emulated physical activity intervention on incident CVD risk, when the physical activity exposure was quantified using self-report vs. wearable devices.

**Methods:** The emulated randomized controlled trial identified physically inactive adults (<150 moderate-to-vigorous physical activity (MVPA) mins/week) who had repeat assessments for wearable and self-reported physical activity. At re-examination, participants were categorised into intervention (adopted the current recommendation of ≥150 MVPA mins/week) or control (remained physically inactive) groups. Participants in each group were propensity score-matched to balance lifestyle behaviours, demographic, and health factors. Cumulative risk for CVD incidence was assessed through cumulative risk curves, hazard ratios, risk ratios, using Fine-Gray subdistribution and Poisson regression models.

**Results:** The wearables analytic sample included 490 participants (245 per arm; mean incident CVD follow-up 4.4 years), and the self-report sample included 11,302 participants (5,651 per arm; mean follow-up 6.3 years). In wearables assessments, guideline-adherent participants had markedly lower cumulative CVD risk (cumulative risk = 8.0% vs. 17.0%; hazard ratio [95%CI] = 0.59 [0.36, 0.98]; relative risk = 0.45 [0.28, 0.72]). In contrast, self-report assessments showed near-identical risk trajectories for intervention and control groups (cumulative risk = 21.6% vs. 21.2%; hazard ratio = 0.98 [0.89, 1.08]; relative risk = 0.92 [0.84, 1.00]). Matching the self-report sample to the wearables sample for lifestyle, demographic, and health factors confirmed these findings.

**Conclusion:** Reliance on self-reported measures of physical activity in TTE studies may obscure emulated intervention effects due to non-differential misclassification, increasing considerably risk of Type II error. Exposure assessment using wearable devices may be essential for valid causal inference in TTE studies of physical activity and CVD risk. Future TTE studies of physical activity exposures should prioritise objective measurements to avoid biased inferences that could affect public health policy and guidelines.

## Introduction

Randomized controlled trials (RCTs) are widely regarded as the gold standard for causal inference in evaluating intervention effects. By randomly assigning participants to treatment or control groups, RCTs minimise differences between study arms aside from the intervention itself, thus allowing any observed differences in outcomes to be attributed causally to the treatment. However, the logistics, costs, and ethical concerns associated with extended follow-up, such as hospitalization and mortality, often limit the feasibility of traditional RCTs^1 2^. Currently, physical activity guidelines are developed using almost exclusively observational prospective studies, the large majority of which used self-reported measures to quantify physical activity^3-5^.

In scenarios where RCTs cannot be practically conducted, target trial emulation offers a rigorous framework for causal inference based on observational data^6^. This approach involves explicitly designing an observational analysis to mirror the protocol of a hypothetical RCT that addresses the causal research question at hand. The goal is to preserve the internal validity of RCTs while leveraging real-world data, thereby enhancing transparency and mitigating key biases often associated with observational research^7^.

Target trial emulation encompasses a two-step process^8^. First, investigators specify a detailed protocol (including eligibility criteria, treatment strategies, treatment assignment procedures, timing of follow-up, outcomes, causal estimands, and analysis plans) that defines the ideal RCT to answer the research question. Second, the components of this protocol are then applied to observational data: eligible individuals are identified, treatment strategies are assigned, follow-up and censoring periods are delineated, and analytic techniques (such as propensity score matching or inverse probability of treatment weighting) are employed to minimise baseline differences in potential confounders between groups except for the treatment.

While target trial emulation designs have gained momentum in recent years^9^ and are advancing observational population study designs^10-13^ by systematically addressing biases that contribute to Type I error (false statistically significant effects). They remain vulnerable to Type II error (false statistically non-significant effects) when the intervention target variable is measured imprecisely. For target trial emulation studies to yield valid causal inferences, accurate and unbiased measurement of the intervention is crucial^14^. This concern is especially pronounced in observational physical activity research, where reliance on self-reported data (e.g. questionnaires) introduces substantial reporting bias and non-differential misclassification error^15^. Self-reported measures of physical activity capture partial behavioural accounts (mostly leisure-time activities lasting for at least 10-15 minutes) with low precision^16-18^. When such misclassification occurs, target trial emulation frameworks may be prone to Type II error, which may artificially nullify findings despite true benefits of physical activity. It is unclear if the markedly low effects sizes reported by the early application of TTE designs in self-reported physical activity research^19-21^ are signs of such Type II error. In contrast, wearable devices are not susceptible to self-report bias and when matched with high resolution machine learning based methods provide greater accuracy in quantifying physical activity continuously with high measurement granularity^22 23^.

We used a target trial framework to mirror an RCT among physically inactive adults that estimated the effects of increasing physical activity to meet guidelines^3^ versus remaining physically inactive on incident CVD risk. To evaluate the impact of measurement type on causal inference estimates, we compared causal effects on CVD risk from self-report and wearables-based physical activity.

## Methods

Comprehensive descriptions of the UK Biobank design have been published previously ^24^. In summary, between 2006 and 2010, about 500,000 adults aged 40– 69 years from England, Scotland, and Wales were enrolled in the large-scale prospective cohort (overall response rate 5.5%).

### Physical activity measurement

#### Self-reports

Baseline self-reported physical activity was measured between 2006-2010 when participants underwent a touchscreen-based short-form International Physical Activity Questionnaire (IPAQ)^25^ assessment. Participants reported their frequency and duration per week of walking, and moderate and vigorous intensity activity lasting for at least 10 continuous minutes^25^. A total of 394,110 participants provided baseline physical activity self-report data. Physical activity was expressed as metabolic equivalent of task (MET) minutes using standard IPAQ scoring procedures, which assign MET values of 3.3 for walking, 4.0 for moderate intensity activity, and 8.0 for vigorous intensity activity. Weekly MET-minutes for each activity type were calculated by multiplying the MET value by the weekly frequency and duration (MET value × days per week × minutes per day) and summed to obtain total physical activity volume. The IPAQ threshold of 600 MET-min/week corresponds to the guideline recommendation of 150 minutes of MVPA per week. Among the participants with baseline physical activity data, 64,382 also had re-examination data.

The exposure window in the TTE analyses was defined as the time from physical activity re-examination (time zero) to CVD incidence or censoring, whichever came first, i.e. for self-report physical activity this began between 2012-2019.

#### Wearables

A subgroup of 103,712 participants later took part in the main accelerometer substudy conducted between 2013 and 2015. Each person was mailed an Axivity AX3 wrist-worn triaxial accelerometer (sampling at 100 Hz) and instructed to wear it continuously on the dominant wrist for seven consecutive days.

For wearables-based physical activity, the exposure window began between 2017-2019, when 5,063 individuals were invited to repeat the accelerometer assessments^26^, using a stratified sampling by sex and 10-year age bands. Participants were scheduled to complete four monitoring sessions approximately three months apart, with flexibility allowing missed early sessions to be compensated later. Among those invited, 3,208 individuals (63%) agreed to participate. All data collection procedures were identical to those used in the baseline accelerometry data collection.

For the wearable devices, the acceleration signal was calibrated and non-wear periods were identified according to standard procedures^27^. Monitoring days were considered valid if wear time was greater than 16 hours, and to be included in the analysis participants were required to have at least four valid monitoring days, with at least one of those days being a weekend day. MVPA was assessed using a 2-stage Random Forest physical activity intensity classifier schema (accuracy = 84% and F-score= 0.83)^28^. We have described the 2-stage schema in detail previously^29^, and we have also appended it as **Supplemental Text 1**. In brief, Physical activity intensity was classified with an accelerometer-based activity machine learning scheme covering sedentary, light, moderate, and vigorous intensity physical activity. The 2-stage activity scheme uses features in the raw acceleration signal to identify and quantify time spent in different activity types^30^ and then intensities in 10s windows.

### Target trial emulation sample

#### Framework

A target trial emulation approach enables the use of observational data to emulate a “target” trial (i.e., the RCT that would have been conducted, were it possible to do so), with exposures framed as hypothetical interventions over a predefined treatment/exposure period^8^. We emulated a target trial in which physically inactive participants at baseline were assigned to one of two groups: an intervention group that met physical activity guidelines at follow-up, or a control group that remained physically inactive and did not meet guidelines at follow-up (**Table 1**). This design conceptualises the “intervention” as the initiation of guideline-concordant physical activity among previously inactive adults.

#### Eligibility

To operationalise this target trial design, we included participants from the UK Biobank cohort who had at least 2 assessments (baseline and follow-up) of wearables or self-report physical activity data. Eligibility was restricted to physically inactive adults at baseline (i.e., not meeting physical activity guidelines at baseline assessment), ensuring that all participants were “entering into” the hypothetical intervention at time zero. Additional information for the target trial emulation design and timeline comparison of the “target trial” and “trial emulation” is provided in **Supplementary Text 2**.

#### Emulated intervention

Participants were classified into the intervention group based on their physical activity status at re-examination (time zero). Those who had adopted sufficient physical activity (≥150 MVPA-minutes/week) were assigned to the intervention group, while those who remained physically inactive were assigned to the control group. This operationalisation mirrors the “target trial” design described in Table 1, where the causal question of interest is whether initiating guideline-concordant physical activity reduces CVD incidence compared with remaining inactive.

### Cardiovascular disease ascertainment

Participant data was linked with electronic health records for mortality through the National Health Service (NHS) Digital of England and Wales or the NHS Central Register and National Records of Scotland up through November 30^th^, 2022. Inpatient hospitalisation data were provided by either the Hospital Episode Statistics for England (October 31^st^, 2022), the Patient Episode Database for Wales (May 31^st^, 2022), or the Scottish Morbidity Record for Scotland (August 31^st^, 2022). CVD was defined as major CVD incidence including ischaemic heart disease, pulmonary heart disease, heart failure, stroke, and hypertensive heart disease.

### Matching participants for lifestyle and health factors

After categorising participants according to whether they met physical activity guidelines (intervention) or not (control), we implemented propensity score matching to balance potential confounding lifestyle and health characteristics between groups. Propensity scores were estimated using a logistic regression model that included the following covariates: age, sex, education, ethnicity, fruit and vegetable consumption, alcohol consumption, smoking status, sleep duration, screen time, medication use (antihypertensive, antidiabetic, and lipid-lowering medication), prevalent cardiovascular disease, prevalent cancer, and parental history of cardiovascular disease or cancer.

To ensure differences in outcomes can more plausibly be attributed to effects of the exposure. This approach simulates the exchangeability of randomised groups by ensuring that, at study entry, the distribution of lifestyle, demographic, and health characteristics is similar between the intervention and control groups. Matching on these lifestyle, demographic, and health characteristics accounts for measured confounding and strengthens causal inference from observational data in the target trial emulation framework. **Supplemental Figure 1** displays a Love plot showing the standardized mean differences for these covariates between the intervention and control groups, both before matching (full sample) and after matching (inclusion of only participants with similar lifestyle, demographic, and health characteristics between the two groups). Based on standard practice, standardised mean differences with a value less than 0.1 is indicative of negligible imbalance^31^.

### Statistical analysis

We used Cox proportional hazard regression models with Fine-Gray subdistribution to address competing risk, which allows a more accurate estimation of the cumulative incidence of the outcome in the presence of a competing event that precludes the occurrence of the event of interest (CVD hospitalisation or death). The proportional hazards assumption was assessed using Schoenfeld residuals and no violations were observed. To mirror the analyses traditionally used in RCT’s, in addition to hazard ratios, we generated cumulative risk curves. We also used modified Poisson regression models with robust standard errors to estimate risk ratios and risk differences. The control group was used as the reference, when applicable. Propensity score matching was performed using nearest neighbour matching with a 1:1 ratio and a caliper of 0.05 standard deviations. Balance between groups was assessed using standardised mean differences. Participants were excluded from analyses if they had missing data.

In a supplemental analysis, we matched the self-report sample to the wearables sample on lifestyle, demographic, and health characteristics to create comparable cohorts. This allowed us to assess whether differences in CVD risk estimates between self-report and wearables-based physical activity measurements were attributable to exposure assessment method rather than differences in sample characteristics. We did further analyses among adults ≥60 years to mirror interventions targeting older populations^32^, and censoring follow up at 2 years to approximate medium term CVD prevention trials^33^.

If significant risk differences were observed, we conducted the following sensitivity analyses to assess the robustness: 1) exclusion of participants with prevalent CVD prior to time-zero, 2) exclusion of participants with an event in the first year of follow-up, 3) using the upper recommendation (300 minutes/week of MVPA) of the physical activity guidelines to delineate our control and intervention groups, 4) using cause-specific hazard model instead of Fine-Gray subdistribution, and 5) using a negative control outcome of deaths or hospitalisations from accidents (excluding cycling, self-harm, and falls incidence), an outcome that does not have an explicit mechanistic link to physical activity to assess residual confounding. We used R 4.2.1 (R Core Team, 2017) to conduct all analyses. This study was reported in accordance with the TARGET guidelines^34^ (**TARGET Supplemental Table**).

## Results

Our analytic sample for the wearables sample included 490 participants (245 in the control and 245 in the intervention group; **Supplemental Figure 2**) with a mean (SD) follow-up time of 4.4 (1.02) years and 63 CVD incidence events. Our self-report-based sample included 11,302 participants (5,651 in the control and 5,651 in the intervention group; **Supplemental Figure 3**) with a mean (SD) follow-up time of 6.3 (2.4) years and 1,642 CVD incidence events. **Table 2** displays the participant characteristics for both analytic samples.

Figure 1 presents the cumulative risk for CVD incidence for the intervention (meeting physical activity guidelines at re-examination) and control (not meeting physical activity guidelines at re-examination) participants. In the wearables assessed physical activity analysis, participants in the control had a higher cumulative risk over 5 years of follow-up compared to the treatment participants. Specifically, at the end of follow-up participants in the control had a cumulative risk of 16.99% [95% CI = 12.83%, 20.95%] and participants in the treatment group had a risk of 8.06% [5.69%, 11.81%]. In contrast, self-reported physical activity showed minimal differentiation in cumulative risk between control and treatment participants. The cumulative risk increased at a similar rate for both groups, with the control group reaching 21.56% [20.10%, 22.99%], and the treatment group reaching 21.16% [19.64%, 22.64%] over 10 years of follow-up **(Table 3)**.

Analyses using the control group as the reference showed that participants in the intervention group **(Table 3)**, as measured by wearables, had a hazard ratio of 0.59 [0.36, 0.98], while those based on self-report had a hazard ratio of 0.98 [0.89, 1.08]. The corresponding relative risk estimates were 0.45 [0.28, 0.72] for wearables and 0.92 [0.84, 1.00] for self-report. The %risk difference was –2.59% [–4.89%, -0.29%] for wearables compared to –1.24% [–3.23% to 0.75%] for self-reported assessment.

In our supplemental analyses, matching lifestyle, demographic, and health characteristics of the self-report sample to the wearables sample showed no difference in cumulative CVD risk between intervention and control groups for the self-report sample (**Supplemental Figure 4**). Descriptive information for these participants is shown in **Supplemental Table 1** alongside the wearables sample. Among adults ≥60 years and in analysis censoring follow up at 2 years, the wearables sample continued to show lower cumulative risk in the intervention group, wider confidence intervals relative to our main analysis, whereas cumulative risk estimates remained similar between intervention and control groups in the self-report sample (**Supplemental Figures 5 and 6**).

Sensitivity analyses in the wearables sample, including excluding events in the first year of follow-up, excluding participants with prevalent CVD, and redefining the intervention using the upper guideline recommendation (300 mins/week of MVPA) showed results that were consistent with the main analysis (**Supplemental Figures 7-9**). Cause-specific hazard models (**Supplemental Figure 10**) showed risk patterns comparable to the primary Fine-Gray analysis, and the negative control outcome **(Supplemental Figure 11)** exhibited risk patterns that differed from the main outcome, suggesting that residual confounding had minimal impact on our observations.

## Discussion

In this target trial emulation of a guideline-based physical activity intervention, we observed markedly different causal estimates depending on how the emulated exposure was measured. When physical activity was quantified using wearable devices, adopting guideline levels of activity was associated with substantially lower cumulative incidence of cardiovascular disease. In contrast, when the same intervention was defined using self-reported physical activity, risk trajectories for intervention and control groups were nearly identical, yielding null or near-null estimates. Our findings suggest that exposure measurement error, rather than lack of a true causal effect, may be a dominant source of bias in target trial emulation studies of physical activity.

Target trial emulation is increasingly used to strengthen causal inference in observational research by explicitly addressing biases such as confounding and immortal time bias^6 35^. However, our results show that even when these sources of Type I error are carefully mitigated, causal inference remains highly vulnerable to Type II error when the emulated intervention is measured imprecisely. In physical activity epidemiology, self-reported instruments are known to capture only partial behavioural domains, rely on long-term recall, and exhibit substantial random error and non-differential misclassification^15 36^. Under these conditions, true intervention effects may be systematically attenuated toward the null, even within well-specified target trial emulation frameworks.

This perspective also helps to provide important context for interpreting previous target trial emulation studies of physical activity. Recent target trial emulations for COPD^37^, cancer^20 21 38^, mental health^39 40^, obesity^10 41^, and quality-of-life^19^ studies have all relied on self-reported physical activity and often reported weak or null associations. Such findings have sometimes been interpreted as evidence that increasing physical activity has limited causal relevance for these endpoints. Our results suggest an alternative, more parsimonious, explanation: imprecision in the exposure’s measurement may dominate the error structure of these analyses. These masked effects are consistent with experimental and mechanistic^42^ evidence from RCT’s with intermediate outcomes^43^, showing increased physical activity improves vascular function, autonomic balance, and cardiometabolic profiles relevant to CVD risk.

By holding the causal framework constant and varying only the exposure assessment method, our study demonstrates that the ability of a target trial emulation to detect a protective effect can depend critically on how the intervention is operationalised. Notably, the null findings we observed with self-reported physical activity persisted despite substantially higher statistical power, indicating that increased sample size and number of events cannot compensate for poor exposure measurement. They also persisted after matching the self-reported sample to the wearables sample on lifestyle, demographic, and health characteristics. In contrast, wearable-derived physical activity, which captures movement continuously across the full intensity spectrum with high temporal resolution, produced clearly separated cumulative risk curves and effect estimates consistent with established biological plausibility^44-47^ and prior evidence^48-51^.

Taken together, our results have direct implications for the evaluation of physical activity guidelines as an intervention concept. As target trial emulation methods are increasingly applied to observational datasets, in situations where traditional RCTs maybe less feasible, to inform clinical and public health policy, there is a risk that reliance on self-reported exposure data may systematically underestimate the benefits of physical activity, potentially contributing to misguided conclusions about its effectiveness. Objective exposure assessment should therefore be viewed not as a methodological refinement but as a prerequisite for valid causal inference in this context.

Strengths of our study include the explicit target trial specification, the use of repeat exposure assessments to emulate intervention adoption, the direct comparison of self-reported and wearable-based physical activity within the same causal framework, and robust outcome ascertainment through linked health records. This framework allowed us to empirically isolate the impact of exposure measurement on causal estimates, an issue that has been widely discussed but rarely demonstrated. The study design, together with the supplemental and sensitivity analyses in the wearables sample, consistently showed risk differences, underscoring the stability of our observed protective association. Limitations of our study include the limited representativeness of the UK Biobank cohort and the smaller sample size and shorter follow-up in the wearable subgroup, although these factors did not prevent detection of clear and clinically meaningful risk differences. We defined intervention status at re-examination, which may not fully capture long-term maintenance of guideline-level activity and could contribute to attenuation of the associations. As with all observational and target trial emulation studies, residual confounding cannot be fully excluded despite extensive matching on lifestyle, demographic, and clinical factors and our negative control analysis. However, any such residual confounding was unlikely to explain the differences between self report and wearables based analyses, since both analyses incorporated the same set of confounders.

## Conclusions

In our target trial emulation, causal estimates for the association between physical activity and cardiovascular disease differed fundamentally according to exposure measurement method. Wearable-derived physical activity revealed robust protective emulated effects, whereas self-reported measures yielded null findings. These results indicate that imprecise exposure assessment can substantially increase the risk of Type II error in target trial emulation studies, obscuring true intervention effects. Future target trial emulations of physical activity should prioritise objective, device-based exposure assessment to enhance valid causal inference and to avoid potentially misleading evidence shaping future physical activity guidelines, public health policy, and clinical practice.

## Data Availability

Data is available upon application to the UK Biobank

https://www.ukbiobank.ac.uk/

## Data Resource

This research was done using the UK Biobank, a major biomedical database, under application 25813. Data is available upon application to the UK Biobank.

## Acknowledgement

The authors thank all of the participants and professionals contributing to the UK Biobank.

