## Supplementary material for "Target trial emulation of physical activity and cardiovascular disease risk: What is impact of the exposure assessment method?": Tables and Figures

**Table 1: Comparison of target trial and emulated trial using UK Biobank data**

|  | **Target trial** | **Trial emulation with UK Biobank** |
| --- | --- | --- |
| **Population/**  **Eligibility criteria** | Middle-age to older adults:  -Who do not meet physical activity guidelines  - With unimpaired physical function  -Who are prepared to participate in a physical activity intervention that will randomise them to an intervention arm to meet physical activity guidelines or to a control arm | Middle-age to older adults:  -Who and do not meet physical activity guidelines  - With unimpaired physical function |
| **Trial arms** | Intervention: physically inactive adults who are allocated to the intervention group to meet physical activity guidelines over a 5 to 12 year period | Intervention: physically inactive adults at baseline who meet physical activity guidelines at re-examination and followed for 5 to 12 year period (referred to hereafter as “intervention group”) |
|  | Control: physically inactive adults who are asked not to change behaviour | Control: physically inactive adults at baseline who do not meet guidelines at re-examination (referred to hereafter as “control group”) |
| **Assignment procedures** | Eligible individuals are randomly assigned to intervention or control groups. Radom assignment would ensure balance in lifestyle behaviours, demographics, and health factors. | Propensity score matching is used to retain participants in the intervention and control groups who are balanced for lifestyle behaviours, demographics, and health factors |
| **Time zero / start of follow-up** | Date of randomisation | Date of re-examination (when physical activity was reassessed) |
| **Follow-up period** | 5 to 12 years of intervention period | 5 to 12 years of intervention period |
| **Outcomes** | Cardiovascular disease incidence (hospitalisation or mortality) | Cardiovascular disease incidence (hospitalisation or mortality) |
| **Outcome ascertainment** | Collected according to trial protocol | Linkage to national hospital and mortality records in England, Wales, and Scotland |
| **Causal contrast** | Intention-to-treat: comparison of groups as randomised, regardless of adherence | Per-protocol: comparison of groups defined by actual PA status at re-examination, with exposure fixed from that point onward |
| **Effect measure** | Cumulative risk ascertained through survival curves, risk ratios, and risk differences | Cumulative risk ascertained through survival curves, risk ratios, and risk differences |

**Table 2: Participant characteristics for wearables and self-reported study samples**

|  | **Wearables** | | **Self-report** | |
| --- | --- | --- | --- | --- |
|  | **Control** | **Intervention** | **Control** | **Intervention** |
| N | 245 | 245 | 5,651 | 5,651 |
| Age, years | 64.4 (9.7) | 64.6 (9.0) | 60.8 (7.5) | 60.8 (7.3) |
| Male (%) | 108 (44.1) | 117 (47.8) | 2,861 (50.6) | 2,843 (50.3) |
| Education (%) |  |  |  |  |
| College | 115 (46.9) | 114 (46.5) | 2,962 (52.4) | 2,969 (52.5) |
| A/AS level | 37 (15.1) | 33 (13.5) | 735 (13.0) | 728 (12.9) |
| O level | 42 (17.1) | 45 (18.4) | 963 (17.0) | 952 (16.8) |
| CSE | 8 (3.3) | 12 (4.9) | 152 (2.7) | 155 (2.7) |
| NVQ/HND/HNC | 11 (4.5) | 12 (4.9) | 321 (5.7) | 323 (5.7) |
| Other | 32 (13.1) | 29 (11.8) | 518 (9.2) | 524 (9.3) |
| Ethnicity; white (%) | 225 (91.8) | 224 (91.4) | 5,380 (95.2) | 5,374 (95.1) |
| Fruits and vegetable servings/day | 7.4 (4.3) | 7.4 (3.9) | 7.4 (4.2) | 7.9 (4.3) |
| Alcohol consumption | 11.9 (13.4) | 11.9 (13.8) | 14.2 (16.6) | 14.2 (15.8) |
| Smoking history (%) |  |  |  |  |
| Never | 142 (58.0) | 149 (60.8) | 3,507 (62.1) | 3,489 (61.7) |
| Previous | 86 (35.1) | 80 (32.7) | 1,938 (34.3) | 1,959 (34.7) |
| Current | 17 (6.9) | 16 (6.5) | 206 (3.6) | 203 (3.6) |
| Sleep duration, hrs/day | 8.2 (1.1) | 8.2 (1.0) | 7.2 (1.0) | 7.2 (1.0) |
| Medication use (%) | 51 (20.8) | 56 (22.9) | 1,390 (24.6) | 1,387 (24.5) |
| Parental history of CVD (%) | 119 (48.6) | 125 (51.0) | 3,293 (58.3) | 3,287 (58.2) |
| Parental history of cancer (%) | 75 (30.6) | 72 (29.4) | 1,854 (32.8) | 1,832 (32.4) |
| Screen time, hrs/day | 3.8 (1.9) | 3.8 (2.1) | 3.9 (2.2) | 3.9 (2.1) |
| Prevalent cancer (%) | 5 (2.0) | 4 (1.6) | 196 (3.5) | 202 (3.6) |
| Prevalent CVD (%) | 13 (5.3) | 11 (4.5) | 462 (8.2) | 485 (8.6) |

Values represent mean (SD) unless noted otherwise

**Table 3:** Cumulative risk, hazard ratio, relative risk, and %risk difference between control and intervention groups in the wearables and self-report samples

|  | **Wearables** | | **Self-report** | |
| --- | --- | --- | --- | --- |
|  | **Control** | **Intervention** | **Control** | **Intervention** |
| **Cumulative risk** | 16.99 [12.83, 20.95] | 8.06 [5.69, 11.81] | 21.56 [20.10, 22.99] | 21.16 [19.64, 22.64] |
| **Hazard Ratio** | Ref | 0.59 [0.36, 0.98] | Ref | 0.98 [0.89, 1.08] |
| **Relative Risk** | Ref | 0.45 [0.28, 0.72] | Ref | 0.92 [0.84, 1.00] |
| **%Risk Difference** | Ref | -2.59 [-4.89, -0.29] | Ref | -1.24 [-3.23, 0.75] |

Cumulative risk represents the risk at the maximum study follow-up time

Sample size: N=245 in both groups for the wearables sample and N= 5,651 in both groups for the self-report sample

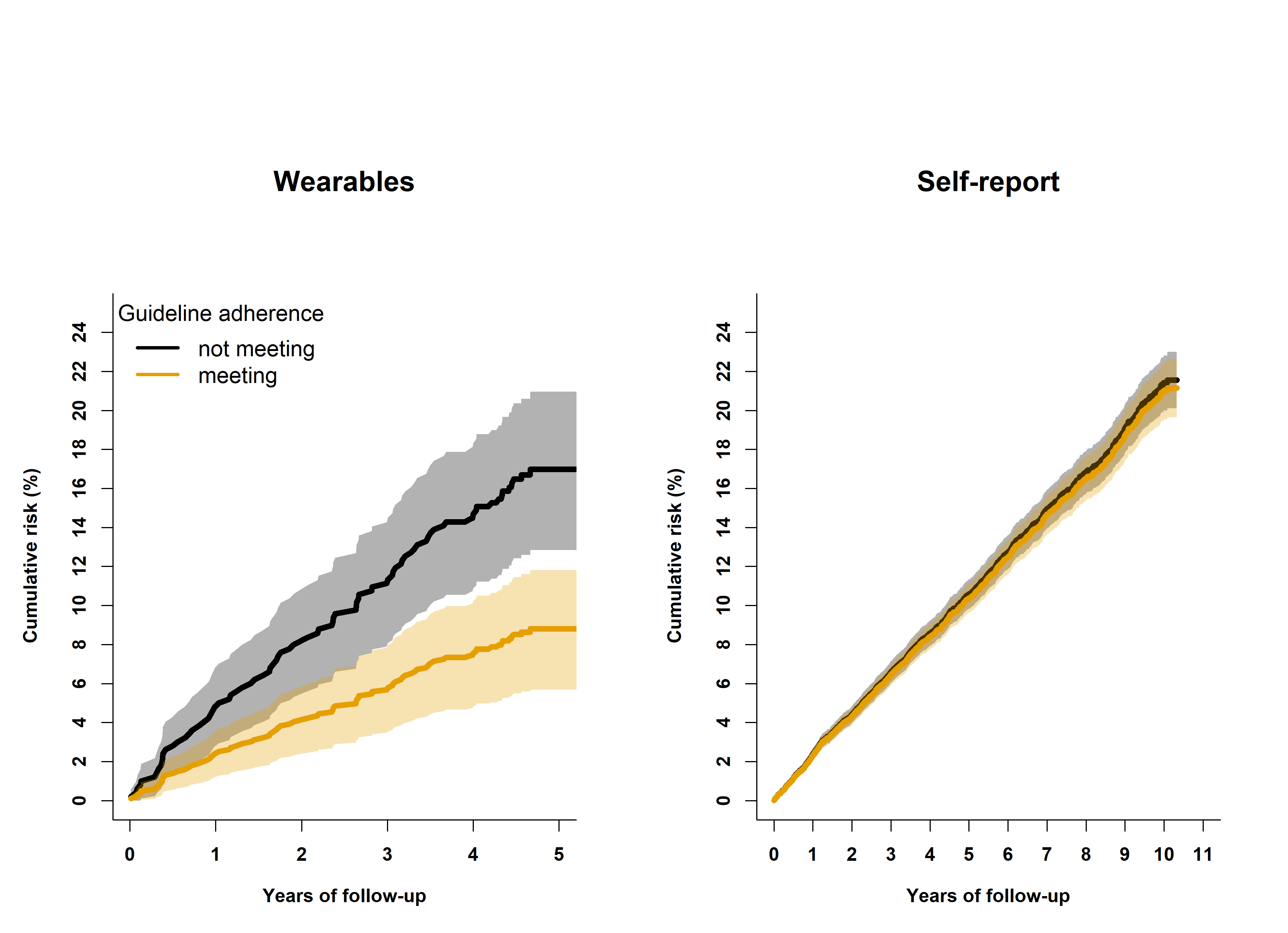
**Figure 1:** Cumulative risk for cardiovascular disease incidence between control and intervention groups when using wearables or self-report to assess physical activity

Participants in the control (not meeting guidelines) and intervention (meeting guidelines) groups were matched for: age, sex, education, ethnicity, smoking history, diet, sleep duration, screen time, medication use, prevalent CVD, prevalent cancer, parental history of CVD, and parental history of cancer.

Sample size: N=245 in both groups for the wearables sample and N= 5,651 in both groups for the self-report sample
